# Interchangeability Between Pleth Variability Index and Internal Jugular Vein Variability Index During Laparoscopic Living Donor Nephrectomy: A Prospective Observational Concordance Study

**DOI:** 10.64898/2026.09.11.26362695

**Authors:** Escarramán Martinez Diego, Sanchez Brito Sebastian, Pérez Lozano Eduardo, Escorza Molina Carla Adelina, Noriega Salas Lorena, Candelaria Olguín Ricardo, Perez Nieto Orlando Rubén, Guerrero Gutierrez Manuel Alberto, Solis Perez Gerardo Alberto, Ignacio Rodriguez Guevara, Galeana Gonzalez Liliana Catalina

## Abstract

**Objective:** This study aimed to evaluate whether PVi and IJV-VI demonstrate sufficient agreement and trending ability to support their clinical interchangeability during laparoscopic living donor nephrectomy.

**Methods:** A prospective observational concordance study was conducted in adult patients undergoing elective laparoscopic living donor nephrectomy under mechanical ventilation. Simultaneous paired measurements of PVi and IJV-VI were obtained every 15 minutes during surgery. Agreement between methods was assessed using repeated-measures Bland–Altman analysis. Trending ability was evaluated using polar plot and four-quadrant plot analyses.

**Results:** Thirty-six patients were included, providing 366 paired intraoperative measurements. Bland–Altman analysis demonstrated a small overall bias of 0.06% (95%CI −0.21 to 0.39), with wide 95% limits of agreement ranging from −5.10% to 5.22% and a global percentage error of 55.96% (95% CI 37.81–74.17). Polar plot analysis showed poor trending ability, with an angular bias of 14.2°, angular dispersion of 117°, and only 34.5% of vectors within the clinically acceptable ±30° zone. Four-quadrant analysis demonstrated an overall concordance rate of 84.7% after applying a central exclusion zone.

**Conclusion:** Although PVi and IJV-VI demonstrated a strong positive association during laparoscopic living donor nephrectomy, agreement and trending ability between methods were limited.

**Main Points:**

- PVi and IJV-VI showed a strong positive association, but this did not translate into adequate agreement.
- Repeated-measures Bland–Altman analysis demonstrated minimal bias but wide limits of agreement and a high percentage error.
- Trending ability between both methods was poor, particularly in polar plot analysis.
- PVi and IJV-VI should not be considered clinically interchangeable during laparoscopic living donor nephrectomy.

## Introduction

Laparoscopic living donor nephrectomy is physiologically challenging because pneumoperitoneum can alter venous return, systemic vascular resistance, renal perfusion, and urine output (1,2). In this setting, avoiding both hypovolemia and fluid overload is clinically relevant, and dynamic hemodynamic indices may support a more individualized approach to perioperative fluid management (3,4).

Stroke volume variation (SVV) and pulse pressure variation (PPV) are well-established dynamic indices for assessing fluid responsiveness during mechanical ventilation (MV), but they require invasive arterial monitoring and, in the case of SVV, pulse contour analysis (5,6). Non-invasive alternatives include the Pleth Variability Index (PVi), derived from the photoplethysmographic waveform, and the internal jugular vein variability index (IJV-VI), obtained using point-of-care ultrasonography (POCUS). Both have shown potential utility for dynamic hemodynamic assessment in mechanically ventilated patients (7–8).

Although PVi and IJV-VI are both influenced by cardiopulmonary interactions during MV. Previous studies have primarily evaluated their association with fluid responsiveness or compared them with other dynamic indices, whereas direct evidence regarding agreement and trending ability between PVi and IJV-VI remains limited (9–10). Therefore, this study aimed to evaluate whether PVi and IJV-VI demonstrate sufficient agreement and trending ability to support their clinical interchangeability during laparoscopic living donor nephrectomy. We hypothesized that both methods would demonstrate clinically acceptable agreement and trending ability under standardized mechanical ventilation.

## Materials and Methods

### Study design

A prospective observational concordance study comparing two non-invasive dynamic hemodynamic monitoring indices was conducted between January 2024 and May 2025 at Centro Médico de Alta Especialidad Dr. Antonio Fraga Mouret, Mexico City, Mexico.. This study was reported according to the STROBE statement (11) and the statistical approach was informed by the COMPARE recommendations for haemodynamic method-comparison studies and adapted to the comparison of two non-invasive dynamic indices (12).

### Ethical considerations

The study protocol was approved by the Local Health Research Committee (CLIS R-2025-3501-095) and registered in the UMIN Clinical Trials Registry (UMIN000058061). The study was conducted in accordance with the principles of the Declaration of Helsinki. Written informed consent was obtained from all participants prior to enrollment, and each participant was assigned an alphanumeric identification code to ensure the confidentiality of personal data.

### Study population and variables

Adult patients (≥18years) of either sex who were scheduled for elective living donor nephrectomy (right or left) were included. Patients with known or newly diagnosed arrhythmias, as well as those in whom technical difficulties prevented measurement of the IJV-VI, were excluded. The variables collected included sex, age (years), body mass index (BMI; kg/m²), side of nephrectomy (right or left), and physical status according to the American Society of Anesthesiologists (ASA) classification. For the purposes of this method-comparison study, PVi and IJV-VI were evaluated as two non-invasive dynamic indices of cardiopulmonary interaction. PVi was considered the comparator method, whereas IJV-VI was evaluated as the POCUS-derived alternative method.

### Anesthetic management and POCUS protocol

PVi monitoring was performed using a MightySat® pulse oximeter (Masimo), whereas POCUS IJV-VI was obtained using a Lanmage Mirror 2 Touch ultrasound system (Shenzhen Lanmage Medical Technology). All ultrasound measurements were performed by an anesthesiology resident trained in neck POCUS. For IJV-VI assessment, a high-frequency linear transducer using the carotid artery preset was positioned in the transverse plane at the level of the internal jugular vein and carotid artery. Once an adequate transverse image of the internal jugular vein was obtained, M-mode was activated with the cursor positioned perpendicular to the vessel. Maximum and minimum internal jugular vein diameters were measured throughout a complete respiratory cycle, and the IJV-VI was calculated according to the formula described by Ma et al. (13): (maximum diameter − minimum diameter) / minimum diameter x 100. PVi and IJV-VI measurements were obtained simultaneously, and each concurrent recording was considered a paired measurement. Measurements were collected at 15-minute intervals throughout the procedure. Given the characteristics of the surgical approach, all measurements were performed with the patient positioned in lateral decubitus for donor nephrectomy (right or left side). For PVi measurement, and according to the methodology described by Başaranoglu et al. (14), the pulse oximeter sensor was placed on the middle finger of the dependent upper limb relative to the side of nephrectomy. PVi values were recorded directly from the monitor as displayed average values at the time of each measurement.

All patients underwent standard intraoperative monitoring according to institutional protocols, including noninvasive blood pressure, pulse oximetry, five-lead electrocardiography, capnography with end-tidal carbon dioxide monitoring (EtCO_2_), and neuromuscular monitoring using train-of-four (TOF). Anesthesia induction was performed using fentanyl (3–5 µg/kg), lidocaine (1–1.5 mg/kg), and cisatracurium (0.1 mg/kg). Anesthesia was maintained with sevoflurane, targeting an end-tidal sevoflurane concentration (EtSevo) between 1.5% and 1.9%. To optimize the performance of dynamic indices (PVi and IJV-VI), MV was standardized according to current recommendations (15): tidal volume 6–8 mL/kg predicted body weight (PBW), positive end-expiratory pressure (PEEP) of 5 cmH_2_O, and respiratory rate adjusted to maintain end-tidal carbon dioxide (EtCO_2_) between 33 and 35 mmHg. In addition, the fraction of inspired oxygen (FiO_2_) was adjusted to maintain peripheral oxygen saturation (SpO_2_) between 94% and 97%. Although pneumoperitoneum may alter the performance of dynamic indices, cardiopulmonary interaction–based parameters such as SVV and PPV remain commonly used for perioperative dynamic hemodynamic assessment. Therefore, given their physiological plausibility, PVi and IJV-VI were selected for comparison in the present study (5, 6). Throughout the procedure, pneumoperitoneum pressure was maintained between 10-13 mmHg.

### Sample size

A total of 36 consecutive patients were included during the study period, yielding 366 paired PVi and IJV-VI measurements. Considering the concordance study design with repeated measurements per patient, the sample size was determined pragmatically based on the number of eligible patients during the recruitment period, the availability of living donor nephrectomy procedures at the study center, and the operational feasibility of obtaining standardized serial ultrasound measurements. Because no prior data were available regarding the expected concordance between PVi and IJV-VI during laparoscopic living donor nephrectomy, no formal sample size calculation based on anticipated limits of agreement was performed. Therefore, the sample size was defined pragmatically; however, the analytical approach was centered on the number of patients rather than solely on the total number of paired measurements, explicitly accounting for within-subject correlation arising from repeated measurements.

### Statistical analysis

Study variables were summarized as median and interquartile range (IQR) for continuous variables and as absolute frequencies and percentages for categorical variables. Comparisons between groups according to the side of nephrectomy (right vs left) were performed using the Mann–Whitney U test for continuous variables and Fisher’s exact test for categorical variables.

To evaluate agreement between PVi and IJV-VI, repeated-measures Bland–Altman analysis was performed, accounting for within-subject correlation arising from multiple observations per patient. The difference between methods was defined as PVi − IJV-VI, and systematic bias was estimated as the mean of these differences. The 95% limits of agreement (LoA) were calculated as bias ± 1.96 times the standard deviation (SD) of the differences using a repeated-measures approach based on a mixed-effects model. This model incorporated the within-subject correlation structure, allowing an unbiased estimation of the variability of the differences and, consequently, of the limits of agreement, as previously recommended (16). 95% confidence intervals (95%CI) for bias, LoA, and percentage error (PE) were estimated using cluster bootstrap resampling at the patient level while preserving the within-subject correlation structure derived from repeated measurements. In each iteration, patients were resampled with replacement, and all corresponding observations for each selected patient were retained. This approach was implemented to obtain robust estimates of uncertainty around concordance parameters, as previously recommended for concordance studies with repeated measurements (17, 18).

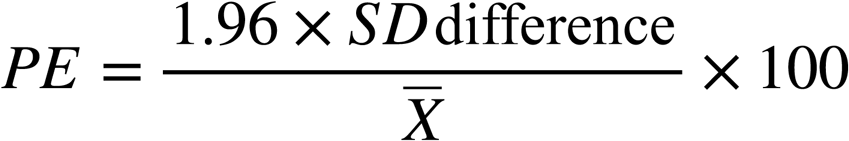

Where SD_difference_ corresponds to the standard deviation of the differences between both methods and ^X^ the overall mean of the measurements. This parameter was used as a complementary descriptive measure to contextualize the relative dispersion between methods. Because conventional PE thresholds were originally developed for cardiac output method-comparison studies, no universal cut-off was assumed to define clinical interchangeability for PVi and IJV-VI. Instead, agreement was interpreted considering bias, LoA, PE, and trending performance collectively. The distribution of the differences and the potential presence of proportional bias were assessed through visual inspection of Bland–Altman plots, examining the dispersion of the differences relative to the mean of the measurements and evaluating the presence of systematic trends across the observed range of values. Clinical acceptability of concordance was interpreted by considering the magnitude of bias, the LoA, PE and the trending ability between methods.

Because static concordance does not necessarily reflect a method’s ability to detect changes in dynamic indices over time, trending ability was evaluated using polar plot analysis. For this purpose, consecutive changes in each index were calculated from serial measurements and defined as:

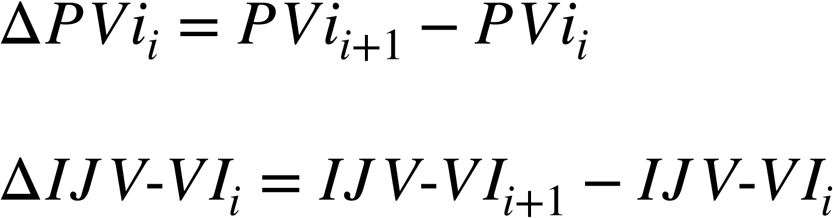

Where ΔPVi*i* and ΔIJV-VI*i* represent the change between two consecutive measurements of the PVi and IJV-VI indices, respectively. Each pair of changes was plotted on a cartesian plane, with ΔIJV-VI represented on the X-axis and ΔPVi on the Y-axis, and subsequently transformed into polar coordinates. The magnitude of the vector was calculated as:

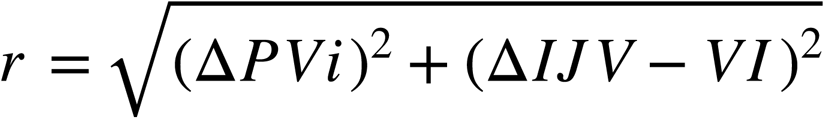

Whereas the polar angle was calculated as:

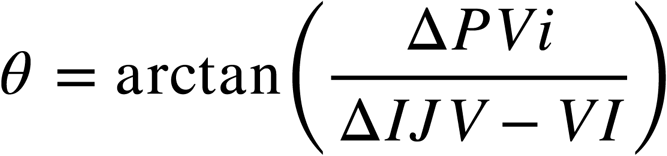

Where arctan denotes the inverse trigonometric tangent function.

For clinical interpretation, angles were adjusted relative to the line of identity (45°), such that a value of 0° represented perfect concordance in the direction of change between methods. Angular bias was estimated as the mean of the corrected angles, whereas angular dispersion was quantified as the standard deviation of these angles. Additionally, the proportion of vectors located within ±30° of the concordance axis (0° after correction) was determined as a criterion for clinically acceptable directional concordance, in accordance with methodological recommendations for polar plot analysis, these thresholds were used as methodological benchmarks derived from haemodynamic method-comparison literature and were not considered universally validated clinical cut-offs for PVi and IJV-VI (19, 20).

As an additional trending analysis, four-quadrant plot analysis was performed, plotting ΔIJV-VI on the X-axis and ΔPVi on the Y-axis. Directional concordance was quantified using the concordance rate, defined as the percentage of pairs of changes located within the concordant quadrants (upper right and lower left). To reduce the influence of physiological noise and low-magnitude variations, a predefined central exclusion zone equivalent to 10% of the mean measurement value was applied, following methodological recommendations for evaluating trending ability using four-quadrant plot analysis (21). Pairs with absolute changes below this threshold were excluded from the analysis.

As a complementary analysis, the monotonic association between PVi and IJV-VI was assessed using Spearman’s correlation coefficient, with 95%Cl estimated using patient-level cluster bootstrap resampling. This analysis was considered exploratory and was not interpreted as a measure of concordance, since correlation evaluates association rather than agreement between methods. As an exploratory sensitivity analysis, agreement, trending ability, and association analyses were repeated after stratification according to the side of nephrectomy. These subgroup analyses were considered hypothesis generating and were not used to modify the interpretation of the primary overall analysis. Graphical representations included Bland–Altman plots, polar plots, and four-quadrant plots. All statistical analyses were performed using RStudio version 2025.09.1+401.

## Results

Thirty-six living kidney donors were included from a total of 39 patients assessed; three patients were excluded due to an ASA physical status ≥ III. A total of 366 paired serial intraoperative measurements were obtained. Median age was 46 years (IQR: 36–55), with a predominance of female patients (63.9%). Median BMI was 26.2 kg/m^2^ (IQR: 23.74–28.5). Most patients were classified as ASA II (91.7%). Regarding surgical positioning, 66.7% of procedures were performed in the right lateral decubitus position. Median PVi was 8% (IQR: 6–11), whereas median IJV-VI was 8.65% (IQR: 5.78–11.44). Additional baseline and perioperative characteristics are presented in Table 1. The dynamic indices evaluated showed comparable values between groups. Median PVi was 9% (IQR: 6–12) in left donor nephrectomy and 8% (IQR: 6–11) in right donor nephrectomy (p=0.12). Similarly, median IJV-VI values were comparable between groups, with values of 8.8% (IQR: 5.45–11.84) in left nephrectomy and 8.5% (IQR: 5.82–11.34) in right nephrectomy (p = 0.80). The remaining between-group comparisons are summarized in Table 2.

**Table 1.**
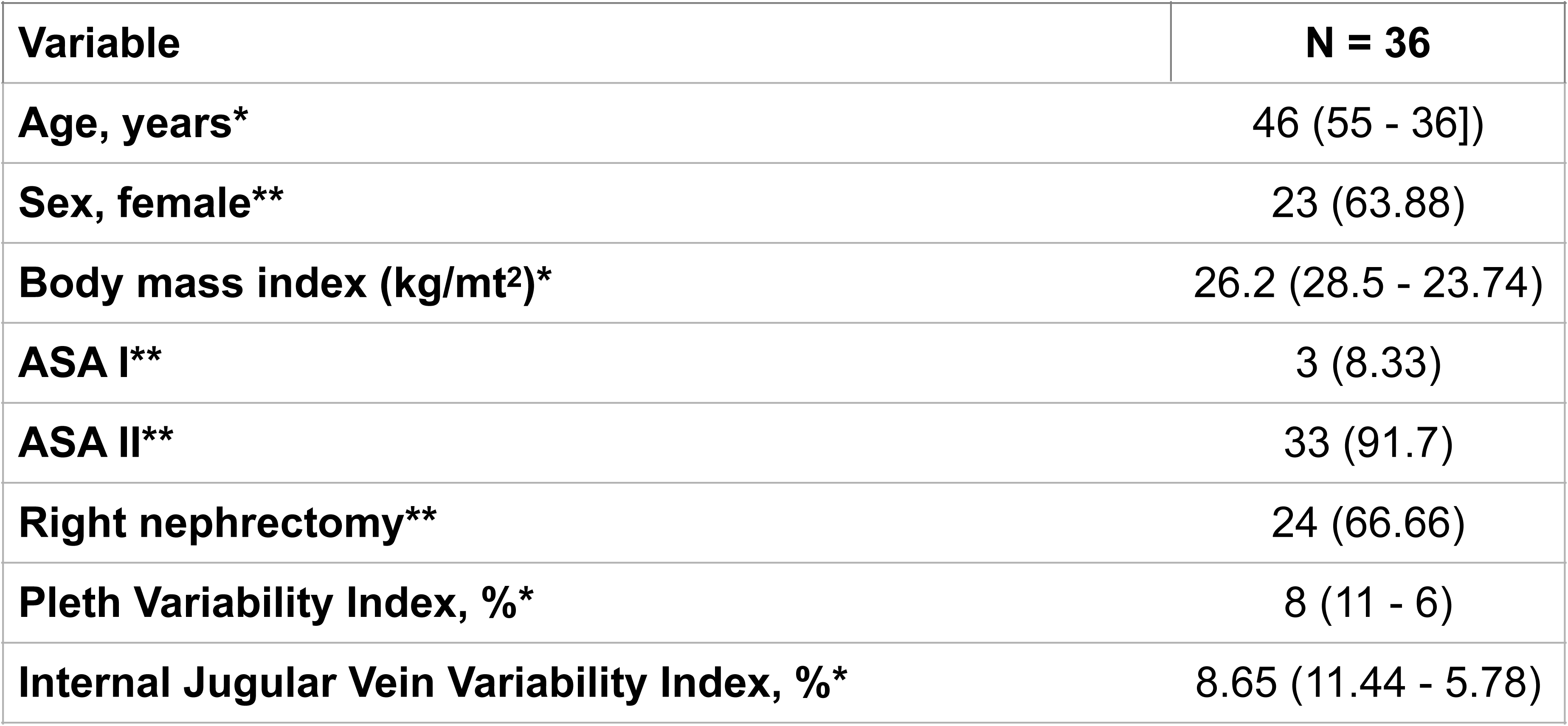
General description of study variables: *Median (interquartile range [IQR]); **Frequencies (%). ASA: American Society of Anesthesiologists.

| **Variable** | **N = 36** |
| --- | --- |
| **Age, years\*** | 46 (55 - 36]) |
| **Sex, female\*\*** | 23 (63.88) |
| **Body mass index (kg/mt<sup>2</sup>)\*** | 26.2 (28.5 - 23.74) |
| **ASA I\*\*** | 3 (8.33) |
| **ASA II\*\*** | 33 (91.7) |
| **Right nephrectomy\*\*** | 24 (66.66) |
| **Pleth Variability Index, %\*** | 8 (11 - 6) |
| **Internal Jugular Vein Variability Index, %\*** | 8.65 (11.44 - 5.78) |

**Table 2.** Comparison between groups according to nephrectomy side. : BMI: body mass index; ASA: American Society of Anesthesiologists; PVi: Pleth Variability Index; IJV-VI: Internal Jugular Vein Variability Index.

| **Variable** | **Right nephrectomy**<br>**( n = 24)** | **Left nephrectomy**<br>**(n = 12)** | **P Value** |
| --- | --- | --- | --- |
| **Age, years** | 45 (36.5-52) | 48 (36-55) | 0.42 |
| **Sex, female** | 16 (66.66%) | 7 (58.3%) | 0.13 |
| **BMI, Kg/mt<sup>2</sup>** | 26.2 (23.4 -28.4) | 26.4 (24.6 - 29.1) | 0.51 |
| **ASA I** | 1 (4.16) | 2 (16.6) | 0.24 |
| **ASA II** | 23 (95.84) | 10 (83.4) | 0.25 |
| **PVi, %** | 8 (11-6) | 9 (12-6) | 0.12 |
| **IJV-VI, %** | 8.5 % (5.82 - 11.34) | 8.8 % (5.45 - 11.84) | 0.80 |

The repeated-measures Bland–Altman analysis demonstrated, in the overall analysis, a systematic bias of 0.06% (95%CI: −0.21 to 0.39), indicating a minimal average difference between PVi and IJV-VI. The LoA ranged from −5.10% to 5.22%. The lower LoA ranging from −6.80% to −3.48%, whereas the upper showed a ranging from 3.15% to 7.53%. The overall PE was 55.96% (95%CI: 37.81% to 74.17%). In the stratified analysis according to nephrectomy side, the right-sided group demonstrated a bias of −0.08% (95%CI: −0.41 to 0.32), with LoA ranging from −5.31% to 5.15%. The PE for this subgroup was 57.15% (95%CI: 30.68% to 80.34%). In contrast, the left-sided group showed a bias of 0.34% (95%CI: −0.11 to 0.83), with LoA ranging from −4.64% to 5.32%. The PE in this subgroup was 53.36% (95%CI: 35.85% to 66.89%) (Figure 1).

**Figure 1.**
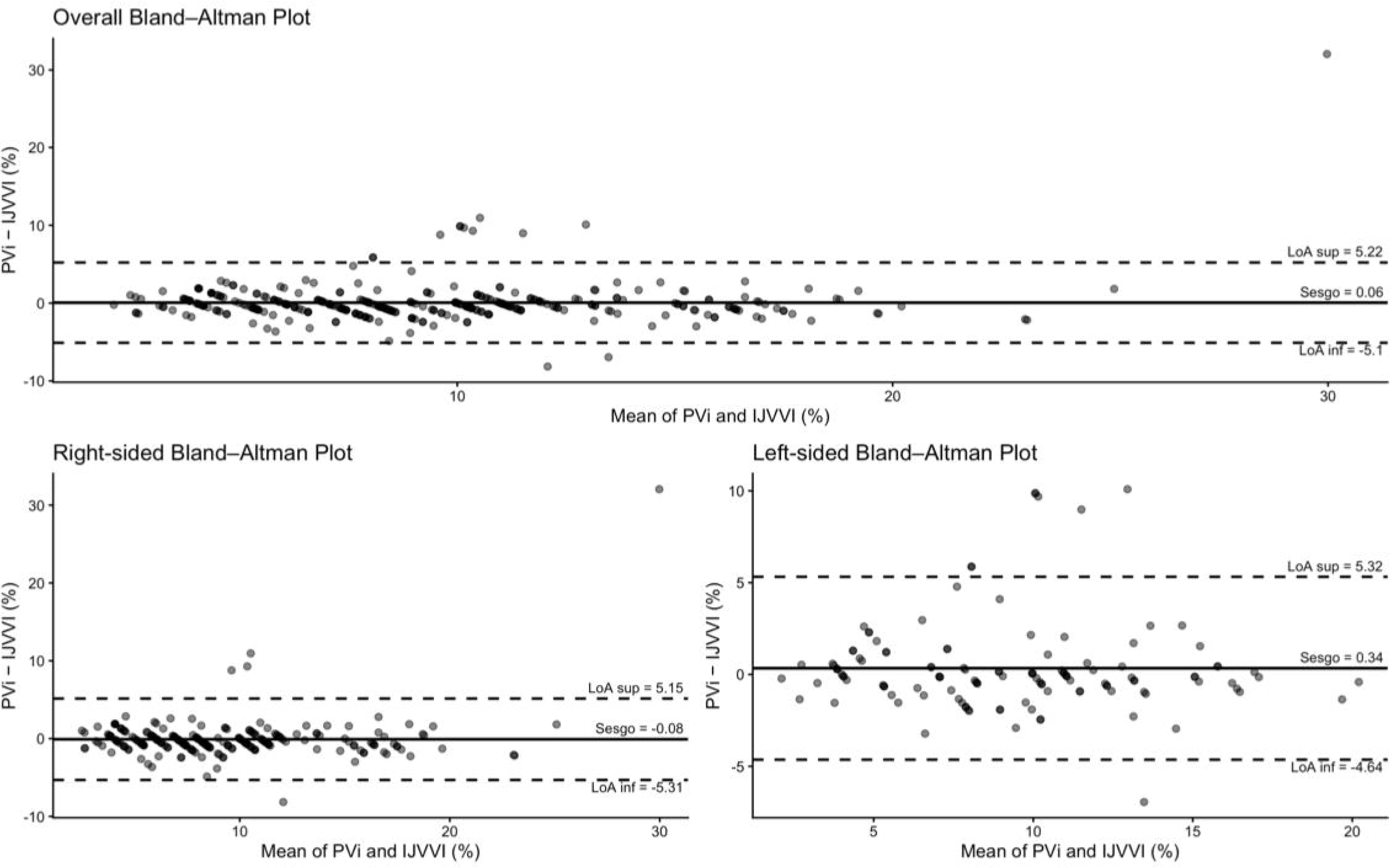
Repeated-measures Bland–Altman plots showing concordance between the Pleth Variability Index (PVi) and the Internal Jugular Vein Variability Index (IJV-VI). Overall and stratified analyses according to nephrectomy side are presented. The X-axis represents the mean of both methods, and the Y-axis represents the difference between measurements (PVi − IJV-VI). The solid line represents the systematic bias, whereas the dashed lines represent the 95% limits of agreement (LoA).

Polar plot analysis demonstrated limited trending ability between PVi and IJV-VI. In the overall analysis, angular bias was 14.2°, with an angular dispersion of 117°, whereas only 34.5% of vectors fell within the clinically acceptable concordance zone of ±30° relative to the corrected line of identity. In the stratified analysis according to nephrectomy side, the right-sided group demonstrated an angular bias of 16.5°, an angular dispersion of 119°, and 35.2% of vectors located within ±30°. In contrast, the left-sided group showed an angular bias of 9.6°, an angular dispersion of 113.2°, with 33.3% concordance within ±30% (Figure 2).

**Figure 2.**
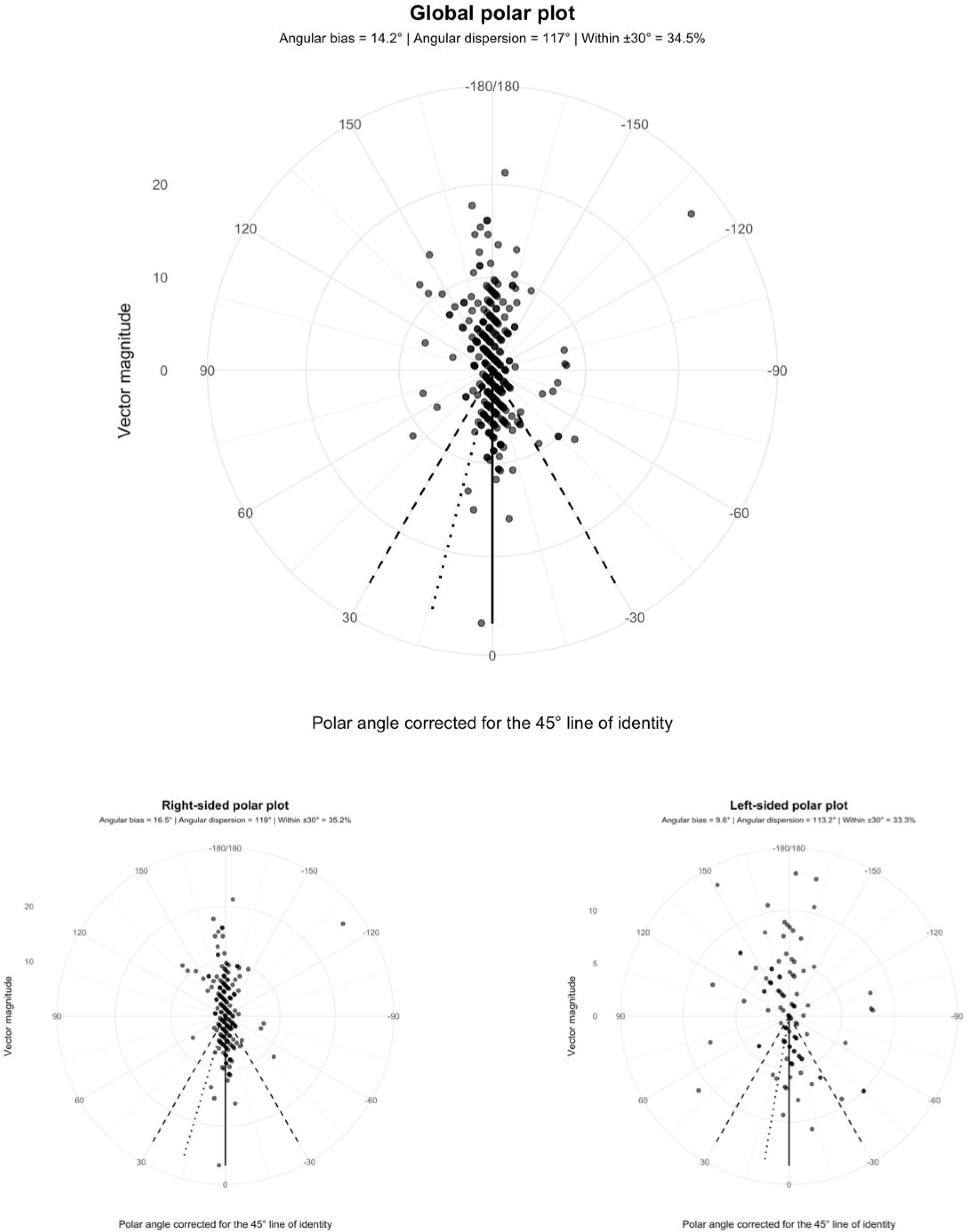
Polar plot analysis for evaluating trending ability between the Pleth Variability Index (PVi) and the Internal Jugular Vein Variability Index (IJV-VI). Overall and stratified analyses according to nephrectomy side are presented. Each point represents a simultaneous sequential change between both methods expressed in polar coordinates, where vector magnitude corresponds to the magnitude of the hemodynamic change and the corrected polar angle represents the degree of directional concordance relative to the 45° line of identity. The solid central line represents the absence of angular bias, whereas the dashed lines delimit the ±30° concordance zone. Angular bias, angular dispersion, and the percentage of vectors contained within ±30° are reported as measures of trending ability.

Four-quadrant plot analysis demonstrated an overall directional concordance of 84.7% after applying a central exclusion zone corresponding to 10% of the mean measurement value (±0.91%). In the stratified analysis according to nephrectomy side, the right-sided group demonstrated a concordance rate of 87.2% using an exclusion zone of ±0.90%, whereas the left-sided group showed a concordance rate of 79.6% with an exclusion zone of ±0.92% (Figure 3). As a complementary association analysis, a strong positive monotonic correlation was observed between PVi and IJV-VI in the overall analysis (Spearman’s ρ=0.876; 95%CI: 0.810–0.925; p<0.001). After stratification according to nephrectomy side, the association was slightly stronger in right-sided nephrectomies (ρ=0.910; 95%CI: 0.861–0.946; p<0.001) compared with left-sided nephrectomies (ρ=0.819; 95%CI: 0.636–0.927; p<0.001).

**Figure 3.**
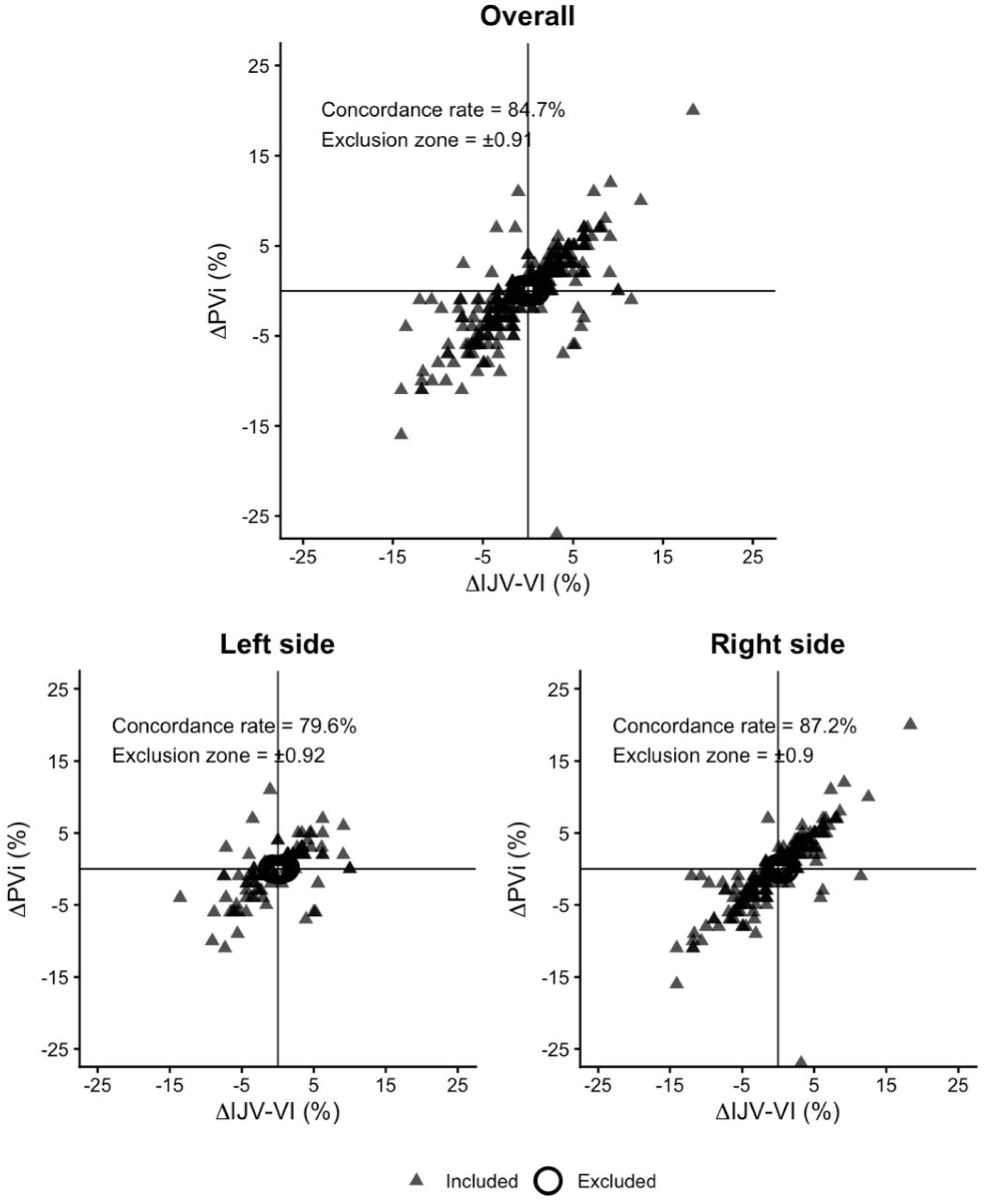
Four-quadrant plot analysis for evaluating trending ability between the Pleth Variability Index (PVi) and the Internal Jugular Vein Variability Index (IJV-VI). Overall and stratified analyses according to nephrectomy side are presented. The X-axis represents sequential changes in IJV-VI (ΔIJV-VI), whereas the Y-axis represents sequential changes in PVi (ΔPVi). Triangles correspond to measurements included in the analysis, whereas circles represent measurements excluded within the exclusion zone. The concordance rate was calculated as the percentage of pairs of changes located within the concordant quadrants, excluding observations falling within the predefined exclusion zone.

## Discussion

In this prospective observational concordance study, PVi and IJV-VI showed a strong positive association; however, agreement and trending ability were insufficient to support clinical interchangeability. Although the mean bias was close to zero, the wide LoA and high relative dispersion indicated substantial variability between methods. In addition, polar plot analysis demonstrated poor trending performance, showing that similar average values did not translate into consistent agreement in either absolute measurements or directional changes over time.

The observed discordance may be explained by the different physiological compartments and biophysical principles underlying each method. Although both indices are influenced by cardiopulmonary interactions during MV, PVi is derived from respiratory variations in the peripheral photoplethysmographic signal and may therefore be affected by changes in digital perfusion and vasomotor tone (22). In contrast, IJV-VI reflects respiratory changes in internal jugular vein diameter and is more directly influenced by intrathoracic pressure transmission, venous return, and central preload dynamics (8). These physiological differences may explain why both methods changed in broadly similar directions while showing insufficient agreement and poor trending ability for clinical interchangeability.

The laparoscopic setting may further contribute to the observed discordance. Pneumoperitoneum alters systemic vascular resistance, venous return, peripheral perfusion, and intrathoracic pressure transmission, potentially affecting PVi and IJV-VI through different mechanisms (23). PVi may be more sensitive to changes in peripheral vasomotor tone and photoplethysmographic signal quality, whereas IJV-VI reflects respiratory variation in a central venous compartment and may therefore be more directly influenced by changes in venous return and preload (8,23). These physiological differences may amplify the discordance between peripheral photoplethysmographic and central venous dynamic measurements. These physiological and biophysical differences may explain why both methods demonstrated a strong association but limited concordance and suboptimal trending ability. Taken together, these findings suggest that both methods may respond in parallel to global hemodynamic changes induced by mechanical ventilation while maintaining differential sensitivity according to the predominant physiological compartment interrogated by each method.

On the other hand, the differences observed according to nephrectomy side, may be explained, at least in part, by physiological and hemodynamic changes related to surgical positioning and pneumoperitoneum during laparoscopic donor nephrectomy. In particular, during left-sided nephrectomies, positioning in the right lateral decubitus position may promote varying degrees of limitation of abdominal venous return and alterations in inferior vena cava hemodynamics, effects that may potentially be accentuated by the increase in intra-abdominal pressure induced by pneumoperitoneum (24, 25). This combination may exert a greater impact on preload and central venous compartment dynamics, leading to alterations in intrathoracic pressure transmission and venous respiratory variations. In this context, IJV-VI may be more influenced by these changes, since this index directly evaluates respiratory variations in internal jugular vein diameter and therefore more closely reflects changes in central and cervical venous return. In contrast, although the lateral decubitus position under general anesthesia has been associated with alterations in ventilation–perfusion matching and changes in respiratory mechanics that may be further exacerbated by pneumoperitoneum (26). PVi may be relatively less affected by these regional mechanical phenomena because of its greater dependence on peripheral vasomotor tone and digital perfusion (22). This may explain why discrepancies between both methods tended to be more pronounced according to nephrectomy side, particularly in scenarios in which alterations in central venous return may be more pronounced.

Previous method-comparison research has also evaluated the interchangeability of ultrasound-derived venous respiratory variation with dynamic arterial indices. Jouffroy et al (27). reported acceptable interchangeability between respiratory variations of the subclavian vein and pulse pressure variation in mechanically ventilated surgical patients, using Bland–Altman analysis. Whether a POCUS-derived jugular venous index can be used interchangeably with a fully non-invasive photoplethysmographic index remains unclear. Fu et al. demonstrated, during major abdominal surgery, a significant correlation between PVi and SVV, as well as adequate predictive performance of both methods for identifying fluid responsiveness (28) Similarly, Pişkin et al. compared PVi with inferior vena cava distensibility in MV patients and observed that both methods demonstrated adequate predictive performance for fluid responsiveness, with similar areas under the curve (29). Regarding IJV-VI, Ma et al. reported that ultrasound-derived internal jugular vein variability was able to predict fluid responsiveness in MV patients after cardiac surgery, showing performance comparable to that of inferior vena cava variability (30). Likewise, Broilo et al. observed that right internal jugular vein distensibility correlated with inferior vena cava distensibility and may behave as a surrogate marker for fluid responsiveness assessment.

These studies primarily focused on association or predictive performance for fluid responsiveness and therefore provide limited information regarding agreement and clinical interchangeability between monitoring methods. In accordance with the principles of method-comparison research, correlation should be interpreted as a measure of association rather than agreement. In the present study, the strong correlation between PVi and IJV-VI did not translate into acceptable agreement or trending ability, underscoring that two methods may track related physiological changes without being clinically interchangeable. These findings suggest that both methods share a common physiological basis related to cardiopulmonary interactions induced by mechanical ventilation; however, they are not necessarily interchangeable in complex scenarios such as laparoscopic donor nephrectomy performed under pneumoperitoneum and lateral decubitus positioning.

This study has several strengths. First, to our knowledge, it is the first to directly evaluate agreement and trending ability between PVi and IJV-VI using a method-comparison approach adapted from the COMPARE framework. Second, the analytical strategy incorporated repeated-measures Bland–Altman, polar plot, four-quadrant, and correlation analyses, allowing assessment of both static agreement and changes over time. Third, ultrasound measurements were obtained using a standardized protocol and recorded simultaneously with PVi during a physiologically complex perioperative setting characterized by mechanical ventilation, pneumoperitoneum, and lateral decubitus positioning. Both monitoring strategies were entirely non-invasive and added no procedural risk. Finally, exploratory stratification by nephrectomy side provided additional physiological context without altering the interpretation of the primary analysis. Limitations should also be mentioned.. First, it was conducted at a single center with a relatively small sample size, which may limit generalizability. Second, IJV-VI is operator dependent, and all ultrasound measurements were obtained by a single trained operator; therefore, interobserver reproducibility could not be assessed. Small variations in transducer pressure may also modify venous diameter. Third, no external physiological reference standard was used because the primary objective was to assess agreement and trending ability rather than diagnostic accuracy for fluid responsiveness. Fourth, pneumoperitoneum, lateral decubitus positioning, and changes in respiratory mechanics may have differentially influenced both methods. Finally, because of the observational design, the proposed physiological mechanisms should be interpreted as plausible hypotheses rather than demonstrated causal relationships.

## Conclusion

Despite a strong positive association, PVi and IJV-VI demonstrated insufficient agreement and poor trending ability, indicating that these non-invasive dynamic indices should not be considered clinically interchangeable during laparoscopic living donor nephrectomy.

## Previous presentation

Not applicable

## Disclosure Statement

The authors have no relevant financial or non-financial interests to disclose.

## Funding

The authors declare that no funds or other support were received during the preparation of this manuscript.

## Ethics Statement

The study protocol was approved by the Local Health Research Committee (CLIS R-2025-3501-095) and registered in the (UMIN000058061). The study was conducted in accordance with the principles of the Declaration of Helsinki.

## Author contributions

DEM: conceptualization and methodology. SBS: original draft and writing. PLE: data curation. EMCA: review and editing, LCGG: data curation and formal analysis. NSL: original draft, writing. CHR: conceptualization. PNOR: review and editing. GGMA: investigation and methodology. SPGA: validation and formal analysis. IRG: writing original draft.

## Data availability statement

The datasets generated and/or analyzed during the current study are not publicly available due to patient confidentiality and privacy considerations but are available from the corresponding author upon reasonable request.

## Notes

### Competing Interest Statement

The authors have declared no competing interest.

### Clinical Trial

UMIN Clinical Trials Registry (UMIN000058061)

### Author Declarations

The Comité Local de Investigación en Salud (CLIS), Hospital de Especialidades "La Raza", Instituto Mexicano del Seguro Social (IMSS), Mexico City, Mexico granted ethical approval for this study. The requirement for informed consent was waived by the Ethics Committee because of the retrospective nature of the study (CLIS R-2025-3501-095).

